# An evaluation of the accuracy of prehospital eFAST in the assessment of polytrauma by a physician-staffed helicopter emergency medical service

**DOI:** 10.1101/2020.12.02.20242453

**Authors:** Christopher Partyka, Matthew Miller, Jimmy Bliss, Brian Burns, Andrew Coggins, Michele Fiorentino, Pierre Goorkiz

**Affiliations:** Aeromedical Operations, NSW Ambulance., Emergency Department, Liverpool Hospital, NSW, Conjoint lecturer, South Western Sydney Clinical School, University of New South Wales; Aeromedical Operations, NSW Ambulance., Conjoint Lecturer, UNSW St George and Sutherland Clinical Schools; Aeromedical Operations, NSW Ambulance, Emergency Department, Liverpool Hospital, NSW, Clinical Lecturer. Sydney Medical School, University of Sydney; GSA-HEMS, NSW Ambulance. Sydney Medical School, University of Sydney; Emergency Department, Westmead Hospital, NSW 2145; Western Clinical School, University of Sydney; Emergency Department, Blacktown Hospital, NSW 2148; Australia; Intensive Care Unit, Liverpool Hospital, NSW 2170. School of Medicine, Western Sydney University

**Keywords:** Ultrasonography, Trauma, Emergency Medical Services, Injury, Focused Assessment with Sonography for Trauma

## Abstract

**BACKGROUND:** While the accuracy of point of care ultrasound in trauma is well understood, there is limited reporting on the efficacy of prehospital ultrasound by helicopter emergency medical service (HEMS). In severe trauma, early diagnosis and communication of life-threatening injuries has the potential to facilitate timely care. This HEMS ultrasound registry evaluation set out to report the accuracy of the extended focused assessment with sonography in trauma (eFAST) exam.

**METHODS:** Retrospective review of trauma patients who received a prehospital eFAST by GSA-HEMS clinicians between 1 January 2013 and 31 December 2017. Clinician interpretations of these scans were compared to immediate in-hospital CT imaging or operating room reports as the gold-standard reference. The primary outcome measure was the accuracy of eFAST for intraperitoneal free fluid compared to hospital CT scan. Secondary outcomes included accuracy of eFAST for pneumothorax, haemothorax and pericardial fluid, comparison of clinician seniority and whether prehospital interventions were supported by eFAST results.

**RESULTS:** We included 896 patients who underwent eFAST by prehospital clinicians. 411 patients had adequate in-hospital data available for comparison. For the primary outcome of IPFF, eFAST had a sensitivity of 25% [95% CI 16-36%] and specificity of 96% [95% CI 93-98%]. Sensitivities and specificities were calculated for pneumothorax (38% and 96% respectively), haemothorax (17% and 97% respectively) and pericardial effusion (17% and 100% respectively). Fifty percent of patients had thoracostomies supported by lung US whilst 24% of patients who received a prehospital blood transfusion had an eFAST negative for haemorrhage.

**CONCLUSION:** This study shows that prehospital eFAST is a reliable tool for ruling in the diagnoses of intraperitoneal free fluid, pneumothorax, haemothorax and pericardial effusion and as expected less reliable than CT imaging for these injuries.

**What is already known about this subject?:**

- Extended Focused Abdominal Sonography in Trauma (eFAST) is widely used in an in hospital setting for the assessment of blunt and penetrating injury.
- Point of care sonography in the prehospital setting has become feasible due to advances in technology, widespread physician training and availability of scanning devices.

**What does this study add?:**

- Our study demonstrates that prehospital eFAST is highly specific for the diagnosis of significant abdominal haemorrhage.
- Prehospital eFAST is less accurate for other injuries including haemothorax and pneumothorax. The explanation for this finding is unclear, but may be associated with scanning earlier in the clinical course, diminishing sensitivity, environmental factors or human factors.
- Further studies are required to understand the optimal role of point of care ultrasound in the prehospital setting.

## INTRODUCTION

Trauma remains a leading cause of morbidity and mortality across both the developed and developing world and thereby systematic improvements in care remain a priority^1^. Improvements in the diagnostic equipment available in the prehospital environment have the potential to expedite the diagnosis of significant injuries, allowing for more targeted immediate interventions whilst facilitating better communication with receiving trauma centres. Unfortunately this may be offset by a diminished accuracy early in the course of the patient journey^2,3^.

Point of care ultrasound (POCUS) has the potential to accurately expedite the diagnosis of critical injuries and thereby improve patient outcomes^4,5^. The extended Focused Assessment with Sonography for Trauma (eFAST) has been shown to be accurate in the diagnosis of abdominal, cardiac and pulmonary injury, both in blunt and penetrating trauma^4,6-7^. This high level of accuracy observed with in-hospital eFAST has not yet translated to the prehospital environment, with recent retrospective HEMS data demonstrating lower sensitivities for both pneumothorax^8^ and intraperitoneal free fluid^9^ detection (28% and 34% respectively). To this end, further knowledge and understanding of prehospital (PH) eFAST has been cited as a high priority in HEMS prehospital care by a number of organisations^10^.

The primary aim of this study was to examine the accuracy of PH eFAST scans for the identification of life-threatening traumatic injuries including intraperitoneal free fluid (IPFF) and pneumothorax (PTx). The secondary aims of the study were to describe the influence of PH eFAST scans on physician clinical interventions and to identify areas for continuing education and training.

## METHODS

### Study Setting

The Greater Sydney Area Helicopter Emergency Medical Service (GSA-HEMS) is a physician-staffed, prehospital and retrieval medicine service treating approximately 1200 injured patients annually across New South Wales (NSW), Australia utilising road ambulances plus rotary and fixed wing aircraft. All patient encounters are recorded in a clinical database (AirMaestro™, Avinet, Australia) which includes demographics, history, interventions performed and clinical observations. Paper-based case-sheets and patient observations are also routinely uploaded to this database. The database contains a dedicated ultrasound registry allowing clinicians to record and store their images, interpretations and report on clinical actions performed as the result of the ultrasound.

This study utilised a retrospective database review of prehospital ultrasounds from the three GSA-HEMS bases where patients were transported to two urban major trauma centres (MTC) in the Sydney Metropolitan area from January 1st 2013 to December 31st 2017. We assessed the clinician interpretation of the scans performed rather than the scans themselves to reflect a real world application of eFAST in the prehospital environment.

### Inclusion Criteria

Patients were included in the study if they met the following inclusion criteria. Firstly, they were identified as having a PH eFAST scan from the AirMaestro database (see Figure 1) and secondly, underwent thoracoabdominal CT imaging or had haemorrhage-control surgery performed within four hours of hospital arrival, were aged>16 and had an ISS>12.

**Figure 1.**
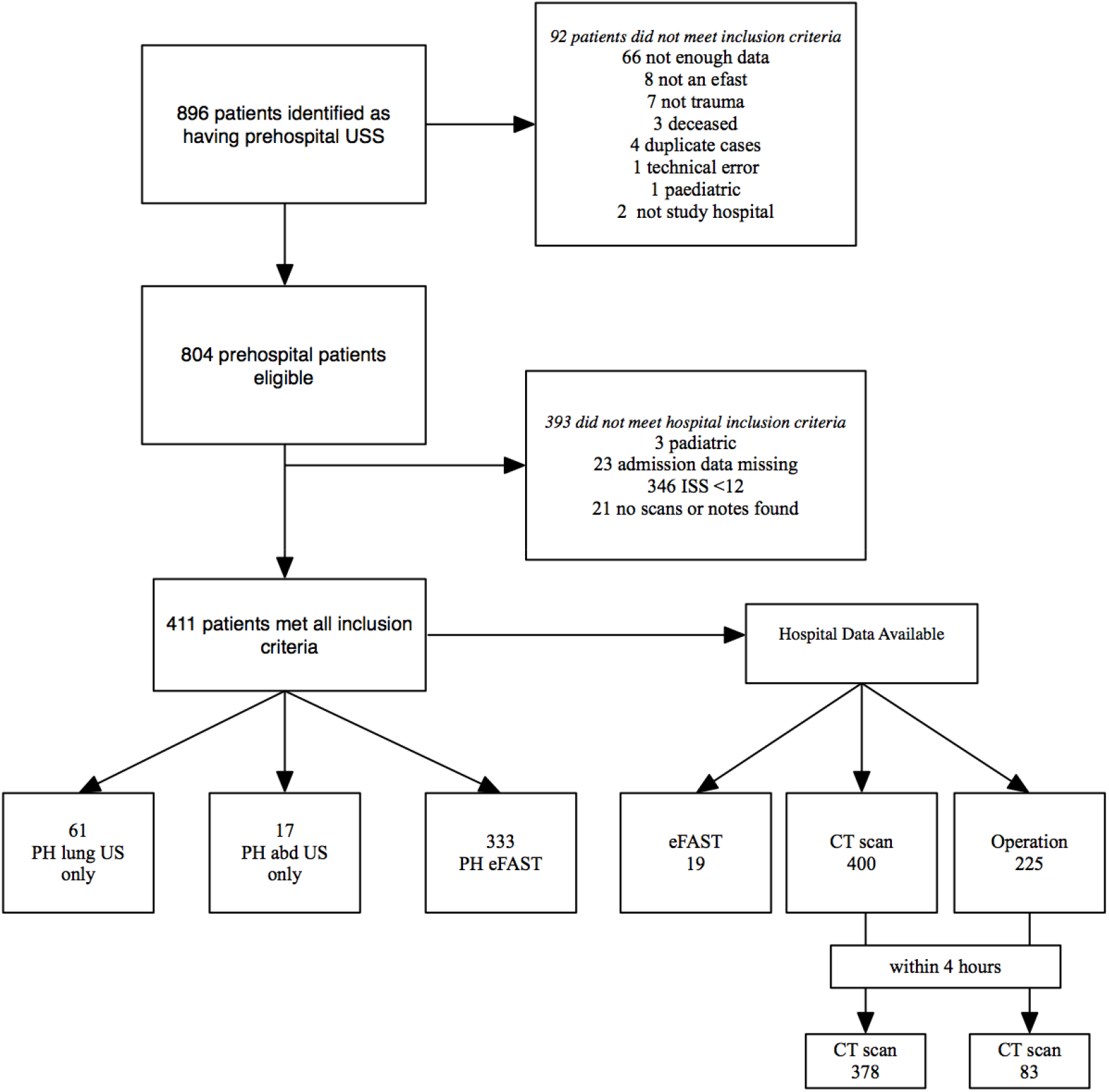
CONSORT diagram.

Prehospital variables were downloaded from the electronic database and cross checked with the patients’ handwritten casesheet for accuracy by two blinded investigators. This included the patient age, gender and arrival date and time at hospital, which were then used to manually identify the patients in the receiving hospitals trauma database. While most study variables could be extracted from this database, the patient’s electronic medical record was also searched for operative, radiology and emergency department admission notes for the gold standard equivalent (either operative findings or CT imaging within four hours of hospital arrival) or Emergency Department eFAST results. The full list of study variables is provided in the supplementary file. PH US can be recorded by physicians as positive, negative or indeterminate if they are unsure. These categories were kept intact in the analysis. All data were entered into a Research Electronic Data Capture (REDCap®) (Nashville, Tennessee, USA) database.

### Outcome Measures

The primary outcome measure was an assessment of the accuracy of PH eFAST interpretation for IPFF. We used either CT imaging of chest/abdomen or intraoperative findings within four hours of hospital arrival as the gold standard investigation. The predefined secondary outcome measures included; (1) assessment of the accuracy of the PH eFAST interpretation for PTx, haemothorax (HTx) and pericardial effusions (2) comparison of PH eFAST interpretation versus in-hospital arrival eFAST by a trauma team member for key diagnoses, where a true positive is presence of the same finding on prehospital and arrival eFAST scan and (3) incidence of prehospital interventions which are performed based on prior ultrasound findings (eg. thoracostomy for detected pneumothorax).

### Power calculation

We performed a power calculation for our primary outcome of assessing PH eFAST accuracy compared to in-hospital eFAST. From previous studies, the expected rate of positive eFAST scans is 10-20%. If we were to detect a difference in positive eFAST of 5% (15-25%) using McNemar test, we would need a sample size of 227 patients at a significance level of 0.95.

### Analysis Plan

Data analysis was performed using the statistical software R (version 4.0, R Core Team; Vienna: Austria) and the epiR package. Parametric or non-parametric summary statistics were produced where appropriate, and 95%CI presented for point estimates. The only hypothesis test planned was McNemar test comparing prehospital ultrasound results with CT scan to diagnose IPFF. In the calculation of diagnostic test accuracy, where no report was found for a finding of IPFF, HTx, PTx or pericardial fluid, it was assumed that the result was negative.

Three post hoc comparisons (not specified in the registered study protocol) were made. Firstly, it is possible that a prehospital ultrasound is initially negative in a patient, but in whom an open thoracostomy is performed, will return a CT positive for pneumothorax as a result of air entrainment into the pleural cavity. We explored this by producing tables of prehospital lung ultrasound against CT results for patients who did, and did not have a thoracostomy. The second post hoc test was to perform a logistic regression of correct prehospital scan against predictors of gender, age, weight, physician grade (consultant/registrar) and mission duration (departing scene to hospital arrival). As this was not prespecified, and two regressions were being performed, we performed a Bonferroni correction to a significance level of 0.025. Finally, we wanted to explore whether a high likelihood of bleeding would change the diagnostic test accuracy for intraperitoneal free fluid. We divided participants into shock index <0.9 and ≥0.9 and repeated the diagnostic test statistics.

This study was approved by our human research ethics committee (LNR/18/POWH/262), and was registered prior to commencement (ACTRN12618001973202) with the original protocol. The main difference between this study and the registered protocol was assessing performance between registrars and consultants by logistic regression, and the post hoc tests mentioned above.

### Patient and Public Involvement

Patients and the public were not involved in the undertaking of this study.

## RESULTS

Overall 896 consecutive patients met the prehospital inclusion criteria of having an eFAST performed during their treatment. Ninety-two were excluded from the prehospital dataset mostly because of incomplete documentation and 393 did not meet in-hospital criteria (Figure 1). Overall, 411 patients were available for data analysis. A missing data analysis is provided as a supplemental file, however 3.2% of data were absent in the final dataset, which was below our prespecified 5% threshold, so complete case analysis was used. There were not enough in-hospital eFAST results to compare with the PH eFAST, the raw results are provided as a supplemental file.

Table 1 presents the demographic data, including those patients who were excluded from analysis for comparison. The majority of the patients were male (73%) and suffered blunt trauma (98%). All groups appear similar in age and gender but those excluded in-hospital may have a lower shock index in keeping with an ISS <12. Of the patients included, 149 were treated by a consultant and 261 by a registrar, 225 (56%) went to the operating theatre at some stage during their admission (for all types of surgery), but only 83 patients were operated on within four hours. For those that had a CT, the median time from arrival at hospital to CT was 66 minutes (IQR 58, 25-75 quantiles 42 to 99 minutes). For those that had a CT within four hours, the median time was 65 minutes (IQR 53, 25-75% quantiles 41 to 94 minutes). The median ISS of included patients was 22 (IQR 12, 25-75% quantiles 17-29).

**Table 1:** Participant Demographics for patients included in the final study sample, and those excluded.

|  | **Included**<br>**( n=411)** | **Excluded prehospitally**<br>**(n=92)** | **Excluded in hospital**<br>**(n= 393)** |
| --- | --- | --- | --- |
| **Age (yrs)**<br>**(median (IQR, 25-75%**<br>**quartiles))** | 39.5(34, 24-58) | 38.5(30, 22-52) | 36(27, 24-51) |
| **Gender (male (%))** | 299 (73%) | 68 (78%) | 285 (73%) |
| **Mechanism:**<br>**Blunt**<br>**Penetrating** | 404 (98%)<br>7 (2%) | 86 (95%)<br>5 (5%) | 365(93%)<br>28 (7%) |
| **Revised Trauma**<br>**Score\*\* (median**<br>**(IQR))** | 7.84 (0.9, 6.9 - 7.8,<br>3.7-7.8) | 7.84 (0.9, 6.9-7.8, 3.1-7.8) | 7.84 (0, 7.84-7.84)* |
| **Shock Index (median**<br>**(IQR, 25-75%**<br>**quartiles))** | 0.79 (0.38, 0.63 -<br>1.00) | 0.77 (0.33, 0.59-92) | 0.69 (0.25, 0.58-0.84) |

*\*\*RTS consistently 7.84 as ISS <12 so observations within the normal limits.*

Table 2 presents the diagnostic statistics for the outcomes of PH IPFF, PTx and HTx. For the primary outcome of PH IPFF, there were 350 PH eFAST scans, 341 with an in-hospital comparison. Four negative prehospital scans and five positive prehospital scans did not have a four-hour hospital comparison, however one of the positive PH eFASTs had hemoperitoneum on laparotomy at 4.5 hours. The sensitivity and specificity of PH eFAST was similar for IPFF and PTx. A comparison of CT results with PH US found CT to be more sensitive than PH US (McNemar test p<0.009).

**Table 2.** Two by two tables with raw numbers followed by diagnostic test criteria (point estimate and 95% CI) for prehospital diagnosis of intraperitoneal free fluid, pneumothorax, haemothorax and pericardial effusion.

| Intraperitoneal Free Fluid |  |  |
| --- | --- | --- |
|  | In hospital CT/ eFAST / Operative findings with four hours of arrival |  |
| **Prehospital US** | *Positive* | *Negative* |
| **positive** | 21 | 9 |
| **negative** | 63 | 257 |
|  |  | Point estimates (95% CI) |
| Apparent prevalence |  | 0.09 (0.06, 0.12) |
| True prevalence |  | 0.25 (0.20, 0.30) |
| Sensitivity |  | 0.25 (0.16, 0.36) |
| Specificity |  | 0.96 (0.93, 0.98) |
| Positive predictive value |  | 0.70 (0.51, 0.85) |
| Negative predictive value |  | 0.80 (0.75, 0.84) |
| Positive likelihood ratio |  | 7.14 (3.40, 14.98) |
| Negative likelihood ratio |  | 0.78 (0.69, 0.88) |
| Pneumothorax |  |  |
|  | In hospital CT |  |
| **Prehospital US** | *Positive* | *Negative* |
| **positive** | 46 | 10 |
| **negative** | 75 | 263 |
|  |  | Point estimates (95% CI) |
| Apparent prevalence |  | 0.15 (0.11, 0.19) |
| True prevalence |  | 0.32 (0.27, 0.36) |
| Sensitivity |  | 0.38 (0.29, 0.47) |
| Specificity |  | 0.96 (0.93, 0.98) |
| Positive predictive value |  | 0.82 (0.70, 0.91) |
| Negative predictive value |  | 0.77 (0.72, 0.82) |
| Positive likelihood ratio |  | 10.00 (5.22, 19.13) |
| Negative likelihood ratio |  | 0.64 (0.56, 0.74) |

| Haemothorax |  |  |
| --- | --- | --- |
|  | In hospital CT |  |
| Prehospital US | Positive | Negative |
| positive | 5 | 6 |
| negative | 24 | 220 |
|  |  | Point estimates (95% CI) |
| Apparent prevalence |  | 0.04 (0.02, 0.08) |
| True prevalence |  | 0.11 (0.08, 0.16) |
| Sensitivity |  | 0.17 (0.06, 0.36) |
| Specificity |  | 0.97 (0.94, 0.99) |
| Positive predictive value |  | 0.45 (0.17, 0.77) |
| Negative predictive value |  | 0.90 (0.86, 0.94) |
| Positive likelihood ratio |  | 6.49 (2.11, 19.95) |
| Negative likelihood ratio |  | 0.85 (0.72, 1.01) |
| Pericardial effusion |  |  |
|  | In hospital CT |  |
| Prehospital US | Positive | Negative |
| positive | 1 | 1 |
| negative | 5 | 276 |
|  |  | Point estimates (95% CI) |
| Apparent prevalence |  | 0.01 (0.00, 0.03) |
| True prevalence |  | 0.02 (0.01, 0.05) |
| Sensitivity |  | 0.17 (0.00, 0.64) |
| Specificity |  | 1.00 (0.98, 1.00) |
| Positive predictive value |  | 0.50 (0.01, 0.99) |
| Negative predictive value |  | 0.98 (0.96, 0.99) |
| Positive likelihood ratio |  | 45.83 (3.23, 649.49) |
| Negative likelihood ratio |  | 0.84 (0.58, 1.20) |

Of PH eFAST, 32% of scans (111 patients) were recorded as indeterminate, of which 27% (30 scans) had either a hospital CT, eFAST or operation finding of positive IPFF. Only 4% of PH lung US were recorded as indeterminate, 53% of which were positive on hospital CT. A total of six patients had hemoperitoneum on operation at greater than four hours, of which four patients had their PH EFAST recorded as indeterminate.

For the outcomes of HTx and pericardial effusion, 394 prehospital lung scans were performed. Three were unilateral (right side only), 255 had a hospital CT, operative note or eFAST for comparison. The sensitivity was 0.20 (95%CI 0.08-0.39) and specificity 0.97 (0.94 to 0.99). Pericardial assessment was reported on 283 prehospital scans performed that also had a CT result for comparison. The sensitivity was 0.17 (95% CI 0.0-0.64) and specificity 1.00 (95%CI 0.98 to 1.00).

To assess if prehospital interventions were supported by their corresponding US scan we reviewed the need for prehospital thoracostomies and blood transfusions. In our study, 60 thoracostomies were performed, 12 had no corresponding lung ultrasound. This meant 48 patients had a prehospital lung ultrasound and a thoracostomy, five of which the ultrasound was reported as indeterminate and for 13 patients, the lung ultrasound was negative for pneumothorax. Therefore only 50% (30 patients) had a thoracostomy supported by a lung US. By comparison, of those without a thoracostomy (351 patients), 302 had a negative PH lung US (86%), 13 (4%) and 28 (8%) were indeterminate or positive. When no thoracostomy had been performed, only 20% of the negative PH lung US demonstrated a pneumothorax. If a thoracostomy had been performed, 84% of the negative PH lung US were reported as having an in-hospital pneumothorax. When we removed the thoracostomy patients who had a negative ultrasound from the prehospital lung US data, the sensitivity and specificity remained similar (supplemental file).

Of the 83 patients who received a prehospital blood transfusion, 20 (24%) had an eFAST negative for either IPFF or HTx. However, three were subsequently found to have mesenteric injury on hospital CT and one had a pelvic fracture. Of the remaining 16 patients, seven went to theatre within four hours and the other nine were lost to follow-up.

The results of the regression models of correct/incorrect scan by patient age, gender, weight, time from scene to CT and physician grade, no variable was a significant predictor for a correct or incorrect scan.

Finally, when we investigated test performance by SI, we found sensitivity was better for high SI patients (0.50, 95% CI 0.23 to 0.73) compared to low shock index (0.32 95% CI 0.17 to 0.51), although their confidence intervals overlap. The positive likelihood ratio was higher for low SI patients (13.2 95% CI 3.9 to 44.5) than high SI (2.6 95% CI 1.4 to 5.4). Sensitivity and negative likelihood ratios were similar (supplemental file).

## DISCUSSION

We performed a retrospective review of the PH eFAST scans performed by our service and compared them to the in-hospital gold standards of in-hospital eFAST, CT imaging or injuries identified at surgery. Our main findings were that the sensitivity and specificity for IPFF, pneumothorax and haemothorax were similar, the caveat being that only a small percentage of the total scans had positive findings. As a result, a low sensitivity translated to a moderate positive predictive value and good likelihood ratios. In addition, we found that, when compared to CT or operative findings, the specificity of eFAST for pneumothorax is high. Finally, in-hospital eFAST reports were documented poorly making the comparison to PH eFAST difficult. Our assessment of pericardial effusion should be interpreted with caution due to the infrequent nature of these cases (2%).

The sensitivities found for IPFF, PTx and HTx were lower than other prehospital studies that used small sample sizes^11-13^, but similar to recent prehospital studies using larger sample sizes for pneumothorax^8^ and eFAST^9^. This highlights the challenge with deploying and interpreting PH eFAST when it is used as a primary assessment tool because the majority of scans are likely to be negative. In other words, scanning every prehospital trauma patient with heterogeneous mechanisms of injury means that, with a larger number of scans performed, and a low prevalence of disease, the negative and positive predictive values are likely to remain low. This occurrence is supported by two meta-analyses demonstrating significant association between prevalence and sensitivity whereby sensitivity falls at a lower disease prevalence^14^. When the likelihood ratios are examined, however, the LR for IPFF and PTx are acceptable^15^. Therefore, when a PTx or IPFF is reported, it is very likely to be present.

The detection of PH IPFF in patients with blunt truncal trauma stratifies that individual to be more likely to deteriorate or need surgical intervention at hospital^16-17^ and should therefore prime the MTC to be ready for such procedures on the patients’ hospital arrival. In multiple patient scenarios, the detection of IPFF should triage the patient for more expeditious transfer to MTC. The higher likelihood ratio for IPFF when the shock index was low, indicates that an eFAST may be of greatest utility in patients in whom overt signs of shock are not yet apparent.

Despite a high specificity, the negative likelihood ratio of PH eFAST suggests caution when interpreting negative studies, as it is still possible to find intra-abdominal blood on CT scan or laparotomy. Our service advises clinicians to treat patients based on their clinical condition, not their scan. We found that 50% of blood transfusions were not informed by PH eFAST. These transfusions may still have been appropriate, as it is possible that intra-abdominal bleeding has not been visualised by ultrasound, or the source of bleeding is external to the peritoneal or pleural cavity. When we reviewed the diagnosis of these patients, over 50% either did have intra-abdominal bleeding or required urgent surgery for another reason. This has implications for quality assurance projects, as when giving prehospital blood, it would be useful if clinicians documented the source of haemorrhage they suspected following patient assessment. Patients with primary head injury frequently receive blood transfusion and have been reported by another service^18^. This may represent a central neurogenic cause of hypotension that is difficult to diagnose in the prehospital multitrauma setting. There may be similar cases in our study although we did not capture these data specifically. We also considered whether false negatives could be due to the accumulation of blood after the PH eFAST, however the proportion of correct and incorrect scans according to prehospital time does not appear to be related to a progression of disease (accumulation of air or blood) in our patients. It is also possible that environmental challenges contribute to the false negative and false positive rates seen in PH eFAST including lighting/brightness, movement of vehicles and imperfect patient positioning. Further exploration into the outcomes of these patients with false negative PH eFAST is warranted.

This study has several limitations. Firstly it examines the clinicians interpretation of the ultrasound images. Whilst this resembles real life practice, the absence of ultrasound storage in our database during the study period prevented independent assessment of the ultrasounds themselves. There is also the possibility of confirmation bias when lung ultrasounds were reviewed as it is feasible that clinicians used ultrasound to confirm a clinical suspicion, for example a pneumothorax and thus recorded/noted a *‘hiss’* once pneumothorax decompressed. This may also be the case with *‘lung down’* finding following thoracostomy and finger insertion into the chest. Patients with chest intervention following a positive eFAST for pneumothorax will often have findings consistent with pneumothorax on CT, potentially confounding the specificity. We examined this by looking at patients with a negative PH ultrasound and reported a much higher false negative rate when thoracostomy had been performed. Removing these scans did not change the test performance, however this may be due to their relatively small numbers. It is important to consider this inflation of false negative rate when performing quality assurance on lung ultrasound.

PH eFAST has different test characteristics to in-hospital scans and we add to the body of literature supporting this. The lower sensitivities reported in this study likely stems from the wider application of PH eFAST as a screening tool used in almost all trauma patient assessments. The performance of interventions despite negative US findings is encouraging as this is in keeping with our Clinical Practice Standard and we do not want US scans to be the only arbiter of treatment judgement. The observation that half of our patients were given blood products without a positive eFAST and appear to have been bleeding on later assessments supports this. High positive likelihood ratios associated with low SI patients highlight the potential for PH eFAST to assist in multiple patient triage scenarios where patients have not yet declared themselves as seriously injured.. Finally, ultrasound quality improvement projects need to ask more defined questions especially in presence of negative findings (for example, why was blood given?, where did you think the patient was bleeding from?, or why was a thoracostomy performed despite a negative PTx scan?), to allow better understanding of US use and its influence on patient treatment.

## Data Availability

Deidentified summary data available if prior ethics approval sought.

## Acknowledgements

We acknowledge Sandra Ware (Research Manager, Aeromedical Operations, NSW Ambulance) and Kait Luker (Database Administrator, Aeromedical Operations, NSW Ambulance) for their work on collating the data set as well as Linda Gutierrez (Trauma Department, Westmead Hospital), Nimmi Kumar and Sally Forrest-Horder (Trauma Department, Liverpool Hospital) for providing access to their local trauma data sets.

